# A Peer-Educator Driven Approach for Sampling Populations at Increased Mpox Risk in the Democratic Republic of the Congo: Implications for Surveillance and Response

**DOI:** 10.1101/2024.10.02.24314817

**Authors:** Sydney Merritt, Megan Halbrook, Yvon Anta, Patrick K. Mukadi, Emmanuel Hasivirwe Vakaniaki, Tavia Bodisa-Matamu, Lygie Lunyanga, Cris Kacita, Jean Paul Kompany, Jean-Claude Makangara-Cigolo, Michel Kenye, Sifa Kavira, Thierry Kalonji, Sylvie Linsuke, Emile Malembi, Daniel Mukadi-Bamuleka, Liliane Sabi, Candice Lemaille, Inaka Marie, Nicola Low, Lisa E. Hensley, Nicole A. Hoff, Robert Shongo, Jason Kindrachuk, Anne W. Rimoin, Placide Mbala-Kingebeni

**Affiliations:** Jonathan and Karin Fielding School of Public Health, Department of Epidemiology, University of California, Los Angeles, CA, USA; Institut National de Recherche Biomedical (INRB), Department of Epidemiology and Global Health, Kinshasa, Kinshasa, Democratic Republic of the Congo; Rodolphe Mérieux Institut National de Recherche Biomedicale Laboratory, Goma, North Kivu, Democratic Republic of the Congo; National Program for Monkeypox and Viral Hemorrhagic Fevers, Ministry of Health, Kinshasa, Kinshasa, Democratic Republic of the Congo; Graduate School for Cellular and Biomedical Sciences, University of Bern, Bern Switzerland; Max Rady College of Medicine, Department of Medical Microbiology & Infectious Diseases, University of Manitoba, Winnipeg, MB, Canada; National Program for HIV/AIDS, Ministry of Health, Kinshasa, Kinshasa, Democratic Republic of the Congo; Institute of Social and Preventive Medicine, University of Bern, Bern, Switzerland; Zoonotic and Emerging Disease Research Unit, National Bio and Agro-Defense Facility, Agricultural Research Service (ARS), United States Department of Agriculture (USDA), Manhattan, Kansas, USA; Department of Internal Medicine, University of Manitoba, Canada; Service de Microbiologie, Département de Biologie Médicale, Cliniques Universitaires de Kinshasa, Université de Kinshasa

**Keywords:** Mpox, MPXV, Democratic Republic of the Congo, Clade I, at-risk populations, sampling strategies

## Abstract

The epidemiological risk factors associated with mpox acquisition and severity in the Democratic Republic of the Congo (DRC) are changing. We assessed perceived mpox risk, and behavioral, clinical and sexual histories among key populations at risk of acquisition through sexual contact. Here, we describe a sampling strategy to enroll participants considered to be at increased risk for mpox infection - men who have sex with men (MSM) and sex workers (SW) - in three urban centers in the DRC. Through the combined approach of time-location sampling with peer educators and respondent-driven sampling, a mixed cohort of 2826 individuals including self-identified MSM (n = 850), SW (n = 815), both MSM and SW (n = 118) and non-MSM, non-SW individuals (n = 1043) was enrolled in Kinshasa, Kinshasa province, Kenge, Kwango province, and Goma, North Kivu province, from March-August 2024. Of these, over 90% were reached through peer educators. The odds of sampling SW individuals were higher at bars/clubs than traditional health facilities. Conversely, the odds of enrolling MSM were highest at selected health facilities. Modifications to the sampling approach were introduced in Kenge and Goma, but these did not affect the enrollment of MSM or SW participants. Ultimately, the selection of, and collaboration with, well-integrated peer educators was the most important facet of this sampling strategy. As the definitions of at-risk populations continue to change for mpox, we demonstrate a functional approach to quickly surveying otherwise hard-to-reach groups for both public health surveillance activities and response.

## INTRODUCTION

The Democratic Republic of the Congo (DRC) was the first country to report human mpox in 1970 and has since experienced the greatest historic burden of the disease. Since 2023, a rapid increase in mpox cases associated with Clade I monkeypox virus (MPXV) in the DRC has been reported, along with geographic expansion within and beyond the country’s borders, including areas previously unaffected by the disease. These events prompted the Africa Center for Disease Control to declare its first public health emergency of continental security and the World Health Organization (WHO) to declare the second public health emergency of international concern for mpox (1, 2).

Viral genome sequencing has demonstrated that APOBEC3 mutations in confirmed mpox cases are correlated with these changing transmission patterns (3–5). As a result, subdivision of Clade I monkeypox virus (MPXV) into Clade Ia (mainly associated with zoonotic transmission) and Clade Ib (associated with sustained human-to-human transmission) have been proposed (3, 6). Most mpox cases in DRC have been mediated primarily through zoonotic transmission of Clade Ia MPXV, with rodents suspected as the primary reservoir. However, the emergence of Clade Ib MPXV has complicated the ongoing public health emergencies due to more sustained human-to-human transmission involving close sexual and non-sexual contacts. This has included cases concentrated within vulnerable populations including sex workers (SW) in eastern DRC (3) as well as sustained transmission via sexual contact among men who have sex with men (MSM) (7). Geographic expansion of mpox cases to urban centers has also been documented, with instances of transmission through sexual contact, including linkages to sex work. Historically, the greatest mpox morbidity and mortality in DRC have been observed in children < 15 years old, while over 90% of Clade IIb mpox infections during the 2022 global outbreak were linked to dense sexual networks, and predominantly among adult males who self-identify as MSM (8). However, the changing patterns of transmission suggest that male and female SW as well as MSM are at increased risk for infection in the DRC.

Sex between men and sex work are not illegal in DRC, but discriminatory regulations and socio-cultural stigma persist, complicating access to these populations and necessitating enhanced sensitivity in outreach and research initiatives (9, 10). National health programs such as the National Program to Control HIV/AIDS and Sexually Transmitted Infections (Programme National de Lutte contre le SIDA; PNLS) include sexually diverse populations in their programming, yet MSM and SW populations are still considered hidden populations in the DRC as there are few demographic data and limited sampling frameworks to reach communities these communities (9, 11). To address these challenges, alternative sampling approaches, such as respondent-driven (RDS) and time-location sampling (TLS), have been proven effective in similar resource limited settings. These approaches have been designed to sample and survey “key populations” who are vulnerable to HIV (12), who face additional barriers in accessing health services, and are otherwise difficult to access (13–15).

Following the 2022 global mpox outbreak, which predominantly spread through sexual contact and disproportionately affected MSM, as well as the documentation of Clade I MPXV transmission through sexual contact in multiple provinces in DRC (7, 16), we sought to survey increasingly at-risk key populations–MSM and SW–using a modified time-location and respondent-driven sampling approach (17).

## METHODS

There is no explicit sampling frame for MSM and SW in the DRC (12). Populations of interest included the following groups: MSM, SW, and a non-MSM, non-SW general population considered at-risk populations (ARP) members at selected venues. ARP members were considered to be at a greater risk than the general DRC population as a whole as they were encountered at locations (such as city entry points, migration centers) and held occupations (such as those at the animal-human interface) where the risk of mpox was thought to be higher than average. Inclusion criteria were adults over 18-years-old; who identified themselves as either men who had ever reported sex with men (regardless of their sexual orientation); participating in sex work; or part of an ARP who voluntary agreement to participate in sampling at selected venues. Participants were excluded from the study if they did not consent to provide a blood sample at the time of interview. Participants were enrolled at sites in three different geographic locations– Kinshasa, Kinshasa Province, Kenge, Kwango Province, and Goma, North Kivu Province. We defined these as “urban centers” due to their high population density, which differs from the rural forested communities, which are more typical mpox transmission sites in the DRC (18).

This sampling approach included six distinct venue types for sampling: bars/cubs, farms or abattoirs, markets, key transition points or points of entry, private residences, and health facilities, including those more accepting for MSM and SW populations. Upon sampling, if the venue did not fall under one of these classifications, it was listed as “other”. Sites were specifically selected in neighborhoods in health zones with known-MSM and SW populations as well as ARPs. In Kinshasa, sites from twelve health zones were included: Gombe, Lingwala, Limete, Kasa-Vubu, Kalamu 1, Kalamu 2, Lemba, Masina 1, Masina 2, Mont Ngafula 1, Kokolo and Nsele. In Goma, sites from three health zones were included: Goma, Karisimbi and Nyirangongo. In Kenge, only sites in the Kenge health zone were included.

### Role of Peer Educators

In March 2024, we recruited and trained peer educators (PEs) who self-identified as members of either MSM or SW populations to direct the community-based enrollment and enroll as “seeds” for our referral-based enrollment strategy. PEs were essential in selecting those venues for TLS as well as recruiting community members through RDS. RDS via PE included the distribution recruitment coupons were labeled such that participants could be linked to the PE recruiter. No financial incentives were provided for PEs to enroll their peers, yet they were paid an hourly wage and provided with communication credits to contact participants via mobile phones. PEs were invaluable resources in the selection of the sampling locations and sensitization of target populations before arrival of the interviewing team. PEs helped to both select TLS venues and seed beginnings of RDS sampling chains – PEs were involved in all aspects of initial participant enrollment.

### Time Location Sampling

The selected sampling method is a flexible, hybrid approach which includes both TLS and RDS to enroll participants for research studies (**Figure 1**) (17). In collaboration with the PNLS, PEs and their established connections with at-risk communities in Kinshasa, Kenge and Goma, we selected specific venues known to be frequented by target groups. These locations included health facilities with specialized care for MSM and SW populations, as well as social venues such as local bars and nightclubs with MSM and SW clientele and key migration points, markets and facilities at the animal-human interface. Sampling at these locations was conducted on a weekly basis at varying times to best recruit and enroll participants - nightlife hotspots were specifically targeted after work hours, to coincide with the most popular hours of operation.

**Figure 1.**
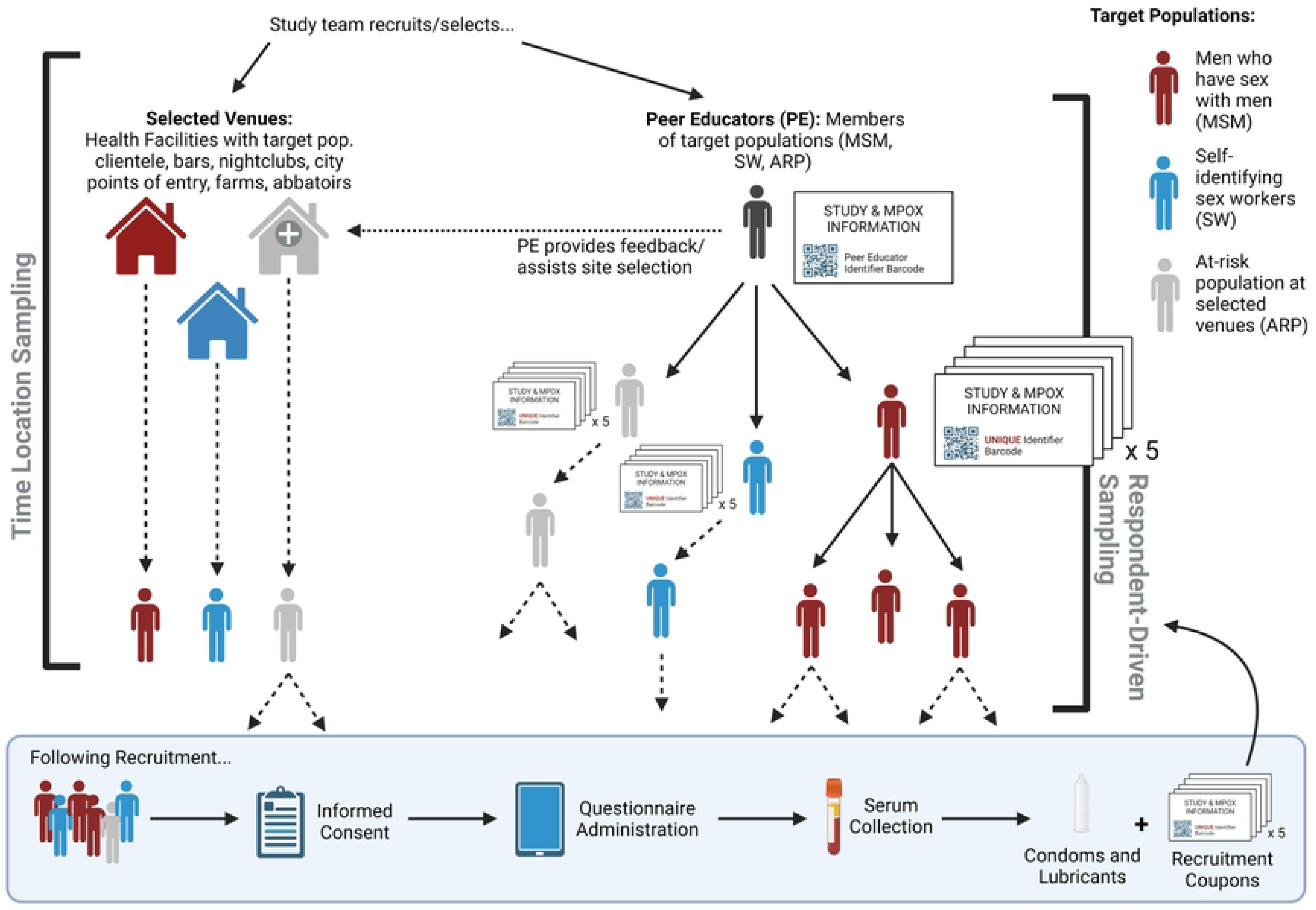
Modified Sampling Approach for the Enrollment of MSM, Sex Workers and At-Risk Population members in Kinshasa, Kenge and Goma, DRC.

### Respondent-Driven Sampling

The study used referral coupons to provide information on how a participant was recruited, either by a peer educator (the initial seed) or another participant, although referral coupons were not required to enroll in the study thus this was a third option. The coupon included information on the study and contact details for the study coordinator, as well as a link to the WHO factsheets on mpox transmission and prevention (in French). On the back of the cards, there was a unique barcode – which indicated how the person was referred and were scanned into the survey at the time of enrollment. Typically, if the person was referred by a peer educator, they would be enrolled at the same time/date and location that they were approached as the study team would already be there or at an organized time/location set by the peer educator. If the person was referred by another participant, they would either enroll on the same date or call the study team to find out the study locations. Upon enrollment at all locations, participants were asked to provide their referral coupon, if available. Following completion of a tablet-based survey, interviewers distributed five referral coupons per participant. Participants were not restricted in who they could share coupons with, but it was assumed most would be shared within their same population of interest.

### At-risk population enrollment

Using similar methods to the MSM and SW population enrollment procedures, ARPs were enrolled at locations considered high risk. At each location, the team would work with a key informant to participate in recruitment of participants – similar to the peer educators in other locations. All potential participants in the ARP were given referral coupons from key informants as well as participant coupons following study completion. All enrolled persons received the same reimbursement within each province.

### Questionnaire Administration and Sample Collection

Trained interviewers interviewed all participants in French or their preferred local language including Lingala, Kikongo and Swahili after obtaining consent. The electronic survey was hosted on a secure survey platform server (SurveyCTO. (2023). *SurveyCTO* (Version 2.0) [Computer software]. Dobility, Inc.). Questionnaire modules included questions related to sexual histories, self-identification of sexual and gender identity, including the ability when asked to define their gender, respondents were given a “select multiple” option, knowledge of mpox, zoonotic and sexual risk factors, clinical symptom history, and general demographic information. Before the interview, a trained phlebotomist at the location took a 9ml venous blood sample in a serum tube and EDTA tube for aliquots of serum, plasma and buffy coat that were stored at -80C for future testing. Post-collection, the interviewer gave the participant a small transportation reimbursement (less than USD3), lubricant, and condoms.

### Data Analysis

Datasets from each urban center were merged to create one complete dataset that was then cleaned and used for additional variable generation. Frequency distributions were calculated for key variables (demographic characteristics, classification as specific populations such as MSM, SW or ARP). Odds ratios were calculated to assess the likelihood of recruiting the target populations at specific venues; “health facility” was set as the reference group as these sites are considered the standard for population serosurveillance recruitment. Kinshasa was solely selected for this model due to the central sampling modification employed in Kenge and Goma. An interaction model joint test was run to assess the interaction between venue type and urban center. All cleaning and analysis were completed in SAS version 9.4 (SAS Institute, Cary, NC).

### Ethical Considerations

This study was approved by the Institutional Review Boards of the University of California, Los Angeles (IRB#23-000676), University of Manitoba (HS25837) and the Kinshasa School of Public Health, University of Kinshasa, DRC (ESP/CE/161/2024). All participants were provided with an informed consent form which was signed or marked in agreement before commencing any study activities. Participant enrollment was conducted from March 20, 2024 to August 25, 2024.

## RESULTS

PEs was the most frequent sampling method, with 96.8%, 99.6% and 98.3% of MSM, SW and MSMSW, respectively enrolled using this approach. PEs almost exclusively recruited all participants - no matter their classification - in Goma (99.1%) and Kenge (98.1%). Convenience sampling was most frequently used among the ARP sample (6.1%) overall and accounted for 17.6% of ARP respondents enrolled in Kinshasa, with 17.6% recruited at the venues themselves. While all participants were provided with five recommendation coupons to seed RDS sampling, only 15 total respondents (0.5%) were recruited using this method **(Table 1)**.

**Table 1.** Recruitment type by target population across all sites.

| Recruitment Type | MSM |  | SW |  | MSMSW |  | ARP |  | All Respondents |  |
| --- | --- | --- | --- | --- | --- | --- | --- | --- | --- | --- |
|  | N | % | N | % | N | % | N | % | N | % |
| **KINSHASA, Kinshasa (n = 940)** |  |  |  |  |  |  |  |  |  |  |
| Peer Educator | 211 | 95.1 | 297 | 99.3 | 105 | 98.1 | 248 | 79.5 | 861 | 91.6 |
| Snowball Sampling | 1 | 0.5 | 1 | 0.3 | 0 | 0.0 | 9 | 2.9 | 11 | 1.2 |
| Convenience Sampling at Venue | 10 | 4.5 | 1 | 0.3 | 2 | 1.9 | 55 | 17.6 | 68 | 7.2 |
| **GOMA, North Kivu (n = 920)** |  |  |  |  |  |  |  |  |  |  |
| Peer Educator | 306 | 99.0 | 203 | 100.0 | 2 | 100.0 | 401 | 98.8 | 912 | 99.1 |
| Snowball Sampling | 2 | 0.7 | 0 | 0.0 | 0 | 0.0 | 0 | 0.0 | 2 | 0.2 |
| Convenience Sampling at Venue | 1 | 0.3 | 0 | 0.0 | 0 | 0.0 | 5 | 1.2 | 6 | 0.7 |
| **KENGE, Kwango (n = 966)** |  |  |  |  |  |  |  |  |  |  |
| Peer Educator | 306 | 95.9 | 312 | 99.7 | 9 | 100.0 | 321 | 98.8 | 948 | 98.1 |
| Snowball Sampling | 1 | 0.3 | 1 | 0.3 | 0 | 0.0 | 0 | 0.0 | 2 | 0.2 |
| Convenience Sampling at Venue | 12 | 3.8 | 0 | 0.0 | 0 | 0.0 | 4 | 1.2 | 16 | 1.7 |
| **ALL SITES (n = 2826)** |  |  |  |  |  |  |  |  |  |  |
| Peer Educator | 823 | 96.8 | 812 | 99.6 | 116 | 98.3 | 970 | 93.0 | 2721 | 96.3 |
| Snowball Sampling | 4 | 0.5 | 2 | 0.3 | 0 | 0.0 | 9 | 0.9 | 15 | 0.5 |
| Convenience Sampling at Venue | 23 | 2.7 | 1 | 0.1 | 2 | 1.7 | 64 | 6.1 | 90 | 3.2 |

Through this modified sampling approach of TLS, peer-educator and respondent-seeded RDS, and convenience sampling of ARP, we enrolled 2,826 participants in Kinshasa (n = 940), Kenge (n = 966) and Goma (n = 920) between March and August 2024.

Of these participants, 850 (30.1%) self-identified as MSM, and 815 (28.8%) self-reported as SW (either as a primary or secondary occupation) **(Table 2)**. Notably, MSM and SW classifications were not mutually exclusive, with 118 respondents (4.2%) identifying as both MSM and SW (MSMSW). Additionally, 1043 participants enrolled were considered part of the general ARP and did not report being an MSM or SW. Most respondents were single (78.6%) and had completed secondary school (42.1%). All respondents reported Kinshasa, Kwango, or North Kivu as their province of residence, apart from one individual (Equateur), indicating that this approach had been successful in specifically targeting urban populations.

**Table 2.**
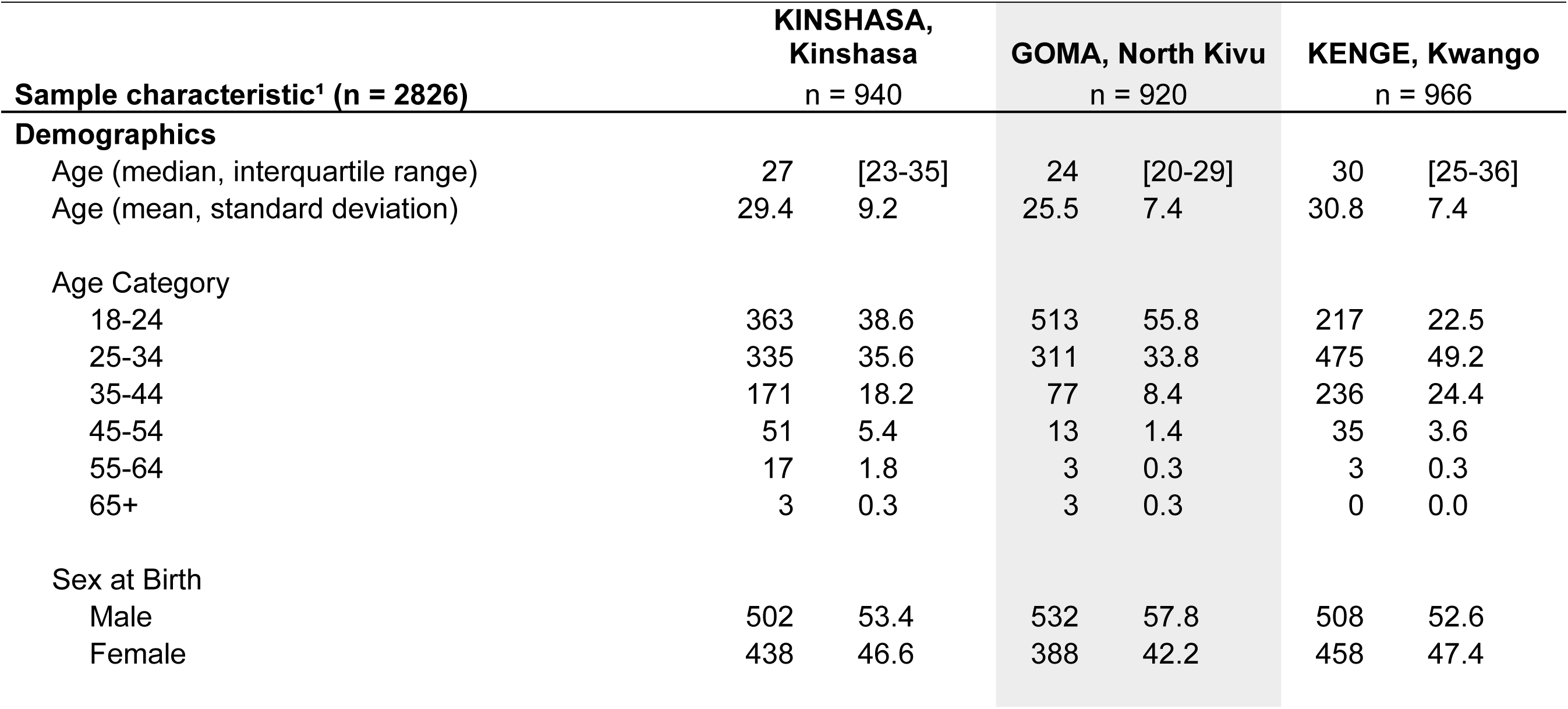
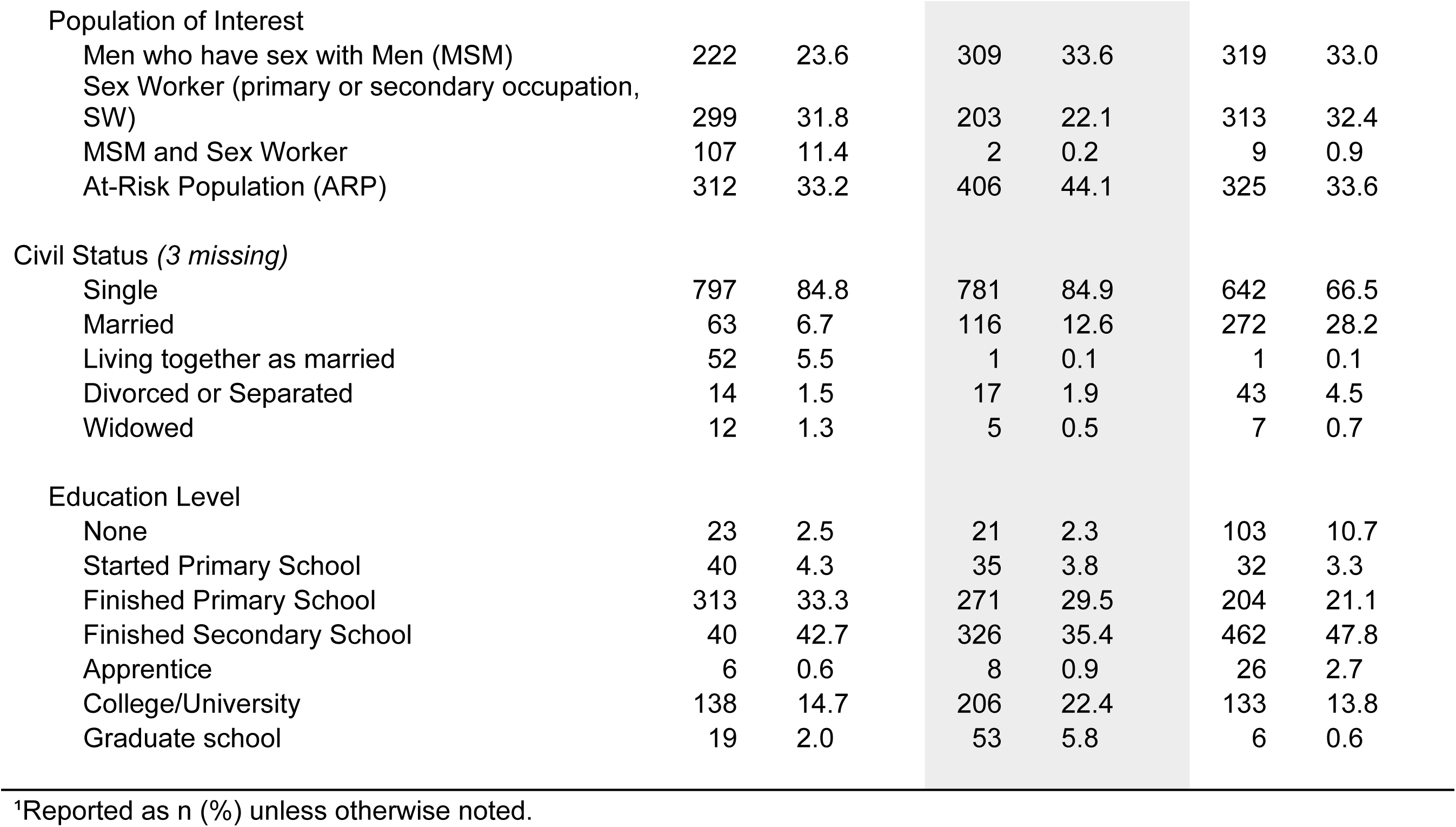
Demographic Characteristics of Urban populations sampled (n = 2826).

We assessed how gender and sexual orientation classifications are understood and the intersection of these self-reported identities (Table 2). No respondents self-identified as queer, intersex or asexual. of note, 2.4% of respondents selected both male/man and female/woman simultaneously (n = 48) or responded with other sex-gender combinations that are incompatible with current definitions **(Table 3)** (19). When dichotomized by sex at birth, male-at-birth respondents reported more sexual orientation diversity than persons responding female-at-birth. Among male respondents, 42.1% identified as straight across all genders. Moreover, 39.4% of male-at-birth respondents self-identified as gay and 18.4% as bisexual. Among MSM-only (defined as male at birth and reporting having sex with men, n = 850), 59.8% of respondents identified as gay, 31.6% as bisexual and the remaining 8.6% identified as straight. Those not classified as MSM overwhelmingly identified as straight (98.8%). Among SW (n = 815), 98.3% of respondents (both male and female) identified as straight. Of the MSMSW population (n = 118), 81.4% identified as gay and 12.7% as bisexual.

**Table 3.**
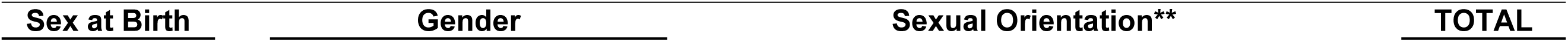
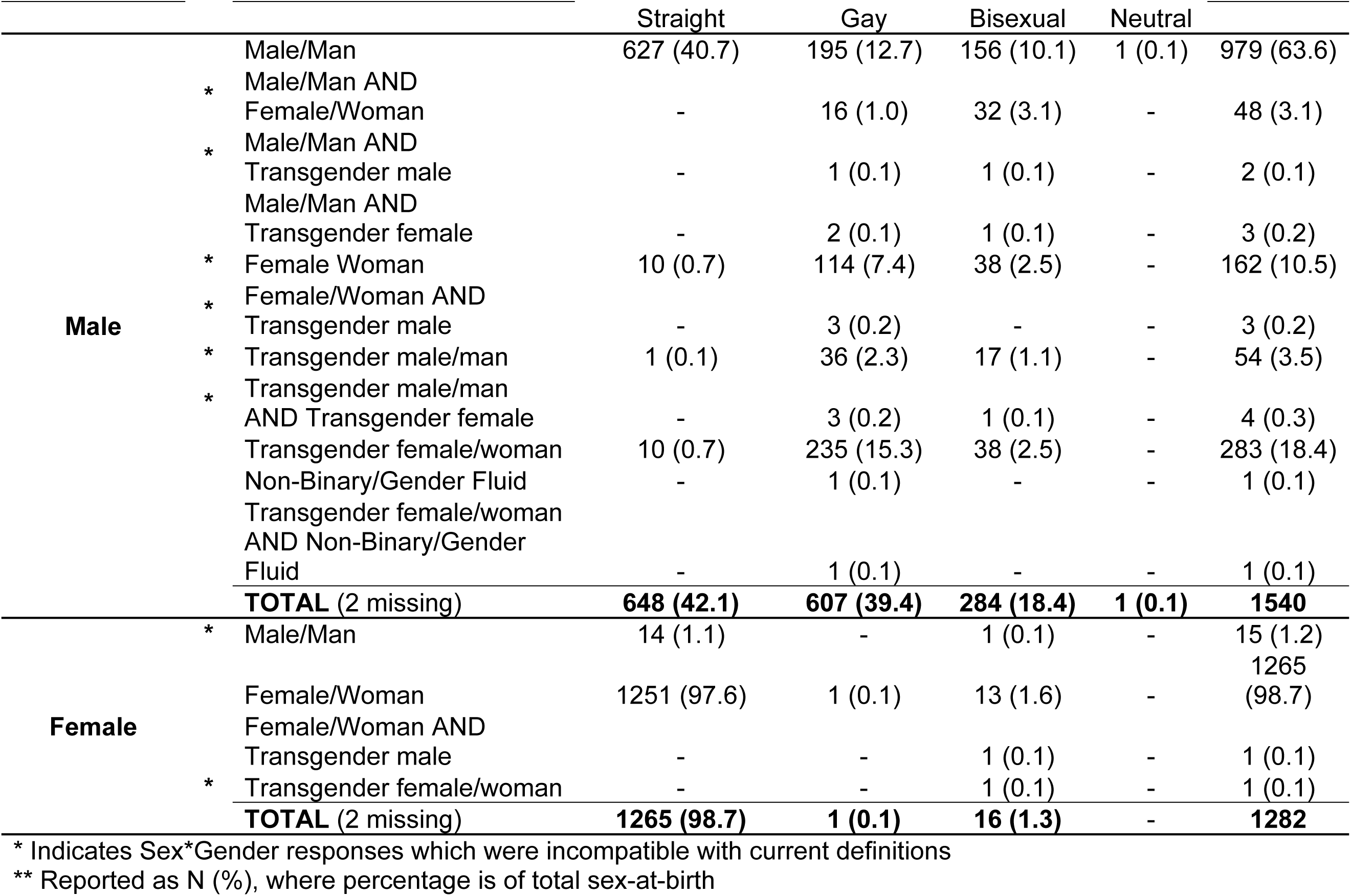
Intersection of self-reported gender and sexual orientation of respondents dichotomized by sex at birth.

| <u>Sex at Birth</u> | <u>Gender</u> | <u>Sexual Orientation**</u> | <u>TOTAL</u> |
| --- | --- | --- | --- |

|  |  | Straight | Gay | Bisexual | Neutral |  |
| --- | --- | --- | --- | --- | --- | --- |
| **Male** | Male/Man | 627 (40.7) | 195 (12.7) | 156 (10.1) | 1 (0.1) | 979 (63.6) |
|  | * Male/Man AND Female/Woman | - | 16 (1.0) | 32 (3.1) | - | 48 (3.1) |
|  | * Male/Man AND Transgender male | - | 1 (0.1) | 1 (0.1) | - | 2 (0.1) |
|  | Male/Man AND Transgender female | - | 2 (0.1) | 1 (0.1) | - | 3 (0.2) |
|  | * Female Woman | 10 (0.7) | 114 (7.4) | 38 (2.5) | - | 162 (10.5) |
|  | * Female/Woman AND Transgender male | - | 3 (0.2) | - | - | 3 (0.2) |
|  | * Transgender male/man | 1 (0.1) | 36 (2.3) | 17 (1.1) | - | 54 (3.5) |
|  | * Transgender male/man AND Transgender female | - | 3 (0.2) | 1 (0.1) | - | 4 (0.3) |
|  | Transgender female/woman | 10 (0.7) | 235 (15.3) | 38 (2.5) | - | 283 (18.4) |
|  | Non-Binary/Gender Fluid | - | 1 (0.1) | - | - | 1 (0.1) |
|  | Transgender female/woman AND Non-Binary/Gender Fluid | - | 1 (0.1) | - | - | 1 (0.1) |
|  | **TOTAL (2 missing)** | **648 (42.1)** | **607 (39.4)** | **284 (18.4)** | **1 (0.1)** | **1540** |
| **Female** | * Male/Man | 14 (1.1) | - | 1 (0.1) | - | 15 (1.2) |
|  | Female/Woman | 1251 (97.6) | 1 (0.1) | 13 (1.6) | - | 1265 (98.7) |
|  | Female/Woman AND Transgender male | - | - | 1 (0.1) | - | 1 (0.1) |
|  | * Transgender female/woman | - | - | 1 (0.1) | - | 1 (0.1) |
|  | **TOTAL (2 missing)** | **1265 (98.7)** | **1 (0.1)** | **16 (1.3)** | - | **1282** |

\* Indicates Sex\*Gender responses which were incompatible with current definitions
\*\* Reported as N (%), where percentage is of total sex-at-birth

In assessing whether the city of enrollment had an impact on the likelihood of sampling for each population, interaction model joint tests were significant for MSM (p <0.001) and MSM/SW (p<0.001), but not SW or ARP (p = 0.67 and 0.097), thus indicating that the role of site type in determining the likelihood of MSM sampling is influenced by urban center of recruitment.

Kinshasa sampling was well distributed across site types, but in both Kenge and Goma, study participants were overwhelmingly sampled at one central location **(Table 4)**. In Goma, sampling was almost exclusively conducted at an “other” facility (88.8%) - the Institut National de Recherche Biomedical, Goma (INRB-Goma). While not a PE selected location, nor a traditional TLS venue for sampling, the INRB-Goma was selected as a central hub for referral of respondents by PE at the selected sites or other participants in response to the ongoing insecurity in the North Kivu region and to ensure safety of staff and participants. PEs referred participants either in person, or via phone calls to known contacts in the community. In Kenge, 97.0% of all respondents were enrolled at a “centre convivial”, a facility with known openness to MSM and SW. Much like the INRB-Goma, one specific facility was selected as a focal point to which all recruited participants were directed. While insecurity is not as great a concern in Kwango province, the central site modification to the sampling approach was chosen to ensure safety of participants who may be targeted due to their MSM or SW identity.

**Table 4.**
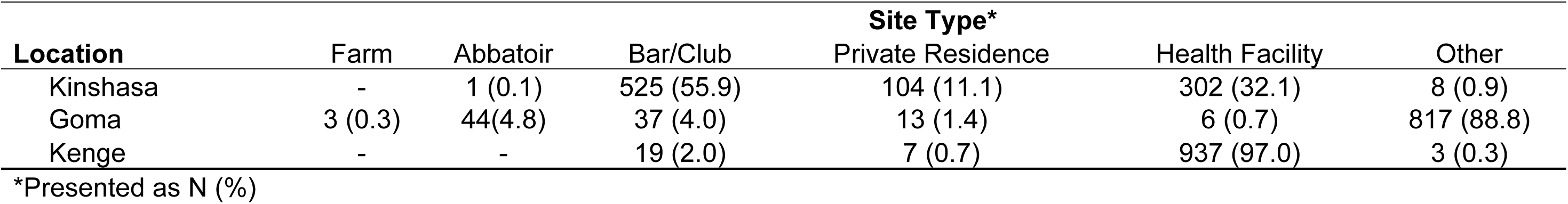
Site Type of participant enrollment by Urban Center Location.

| Location | Site Type* |  |  |  |  |  |
| --- | --- | --- | --- | --- | --- | --- |
|  | Farm | Abattoir | Bar/Club | Private Residence | Health Facility | Other |
| Kinshasa | - | 1 (0.1) | 525 (55.9) | 104 (11.1) | 302 (32.1) | 8 (0.9) |
| Goma | 3 (0.3) | 44(4.8) | 37 (4.0) | 13 (1.4) | 6 (0.7) | 817 (88.8) |
| Kenge | - | - | 19 (2.0) | 7 (0.7) | 937 (97.0) | 3 (0.3) |

\*Presented as N (%)

When looking at the models of the association between venue of sampling and likelihood of recruiting target populations in Kinshasa, the odds of sampling MSM at a bar/club were 1.7 times lower than at a health facility. Using the same parameters, the unadjusted odds of sampling a SW at a bar/club in Kinshasa were 2.8 times higher than at a health facility. MSMSW were similarly 2.4 times less likely to be sampled at a bar/club, and 5.8 times less likely at a private residence than a health facility **(Table 5)**. The odds of sampling general population members were greatest at locations denoted as “private residences” (2.7 times higher than at health facilities). Farm/animal care responses were not included in the model due to low cell counts.

**Table 5.**
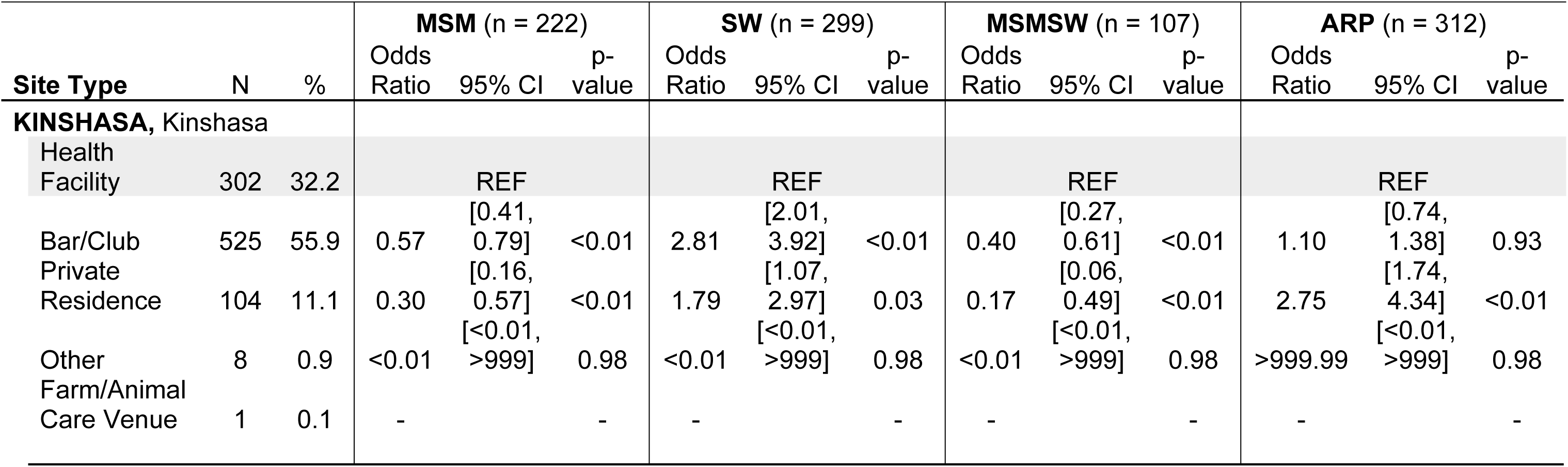
Likelihood of Sampling Target Populations by Site Type in Kinshasa. Farm/Animal venue types were excluded from the Kinshasa-only model due to low cell counts.

## DISCUSSION

In this study, we sampled and enrolled so-called hidden populations, potentially at high risk of acquiring MPXV through sexual contact, in three urban centers in the DRC. Overall, the most effective recruitment strategy was via well-integrated peer educators for MSM and SW target populations. The success of this approach mirrors past HIV-centered care delivery approaches through peer-driven recruitment in similar contexts (20). However, beyond sampling success, we show how, while peer-educators are central to enrollment, sampling venues also play a significant role in sampling likelihood by population type. Additionally, this study expands upon the MSM and SW specific sampling to include general population members considered at higher risk for mpox – a group that has not historically been included in HIV-related MSM and SW population sensitization and sampling (20, 21). Here, we show how a peer-educator driven sampling approach is effective in reaching more than just MSM and SW, but other at-risk groups for mpox, or other diseases transmitted through sexual contact.

In all three locations, RDS sampling accounted for less than 2% of recruited participants. Traditionally, RDS sampling relies on incentives for both recruitment and participation (11) - this may account for the low reported RDS-recruitment. TLS was specifically successful in recruiting SW populations. In Kinshasa, the odds of recruiting SW were consistently higher in selected bar/clubs than a traditional health facility. The opposite was found for MSM, where the odds of sampling this at-risk population were higher at selected health facilities. This finding may be explained by the specific targeting of facilities known as “centres conviviaux”, or those with known tolerance and acceptance of MSM patients. Additionally, these differences observed between MSM and SW recruitment may be attributed to greater socio-cultural stigma for MSM, while SW is an accepted profession in the DRC cultural context (22).

While both MSM and SW populations were successfully recruited in Goma and Kenge, the implementation of the sampling approach was drastically different from that used in Kinshasa. This reliance on PE recruitment may be attributed to the sampling modifications employed in Goma due to security considerations. Additionally, this may highlight gaps in interviewer training– the coupons provided may have not been well explained, or without any incentive, participants may have been less likely to recruit their peers. Yet again, this emphasizes how PEs, even through phone-contact, are essential to this sampling approach. In Kenge, similarly almost all respondents were recruited via PE contact (98.1%) and directed to sampling at a set health center.

The sampling strategy employed in this study was heavily influenced by, and reliant on, peer educators recruited specifically for their connections to the populations of interest. In using PEs for their selection of recruitment venues, as well as their social links to MSM and SW, this sampling design is inherently biased towards successful recruitment by PEs. However, due to the need for rapid data collection, this type of survey is an important mechanism for reaching broad at-risk populations in settings like the DRC.

In a larger low-resource context, public health measures and interventions for these groups should also be developed in conjunction with the groups with which they seek to serve. For example, by integrating peer educators in the study design and implementation, this approach was successful in recruiting both SW and MSM. Community engagement was a central pillar of this study design - the value of well-integrated PEs should be underscored not only through peer recruitment but also through the guided selection of MSM and SW-centered health facilities and venues. Selected health facilities in Kinshasa may have been the best venue for recruiting MSM as they are known to the community as MSM-accepting.

One key aspect of this sampling strategy was defining at-risk populations. Due to the evolving nature of MPXV transmission, MSM and sexually diverse populations were of particular interest for sampling, yet this sensitive information is not readily available. Furthermore, this information may also not be readily shared with interviewers and members outside of the “hidden” population due to fear of persecution. For example, as of June 2024, a new bill has been proposed in DRC which specifically condemns homosexuality and suggests arresting or fining members of these communities (23).

While all interviews were conducted by health-trained professionals, self-reported sex at birth and gender responses were not always compatible with typical definitions of these identifiers (**Table 2**). For example, several respondents reported “male at birth” but also “transgender male”. Additionally, when asked to define their sexual orientation, responses “queer”, “intersex” and “asexual” were never selected, indicating that these terms may either be misunderstood or not used in the DRC context and likely need further piloting to ensure correct terms are being used. Understanding of gender may be linked to socioeconomic status or influenced by cultural or religious contexts. Additionally, given these differing classifications of sex and gender, the MSM population was defined in this context by those respondents who were male at birth. In DRC, gender affirming surgery and hormone therapy are not readily available, thus, those respondents who identified as transgender are not likely reflecting biological status but rather a preferred gender expression. These varying responses of gender and sexual orientation highlight differences in interpretations or potential alternative terms that may be used to best define these populations. As an already hidden population, ascertaining this information is invaluable for expanding definitions of populations who may be at risk for mpox and crafting public health interventions.

One drawback to this sampling approach was the overlap of individuals who were both classified as MSM and SW. Specifically, of 4.2% of respondents (n = 118) identified as MSM and reported sex work as their primary or secondary occupation. This subset of the population is of particular interest for mpox investigation due to potential interacting risk factors. In future analyses including serologic evaluation and establishment of additional mpox risk factors among these communities, these 118 individuals may be assessed separately to better understand MSM and SW-specific risk activities and the role of compound risk. Additionally, another limitation of this approach was lack of venue-specific details to identify where exactly sampling occurred. Specific names of venues were not collected so as to protect the exact whereabouts of these otherwise hard-to-reach populations, yet without this granular information, it is impossible to determine if certain sites were more influential in driving sampling success.

Ultimately, the primary objective of this study was to access and sample key populations to determine if there was evidence of mpox circulation across the DRC. Here, we highlight the peer-educator driven approach as an appropriate strategy for effectively reaching hidden MSM, SW and ARP populations of three urban centers in DRC, largely through the channels of well-integrated peer educators. Given the planned mpox vaccine deployment in DRC in autumn 2024 for at-risk populations, this approach could be expanded to assess general acceptability of mpox vaccines, and other mpox-related interventions. Furthermore, the larger risk factor data and serum samples collected from these 2826 individuals will be analyzed to ascertain perceived mpox risk, and anti-MPXV seroprevalence among these groups.

By accessing these key populations, these approaches could also be used to tailor mpox-prevention and educational strategies. However, when surveying these groups, recruitment and questionnaire material should be framed so as not to additionally discriminate against already stigmatized populations - public health messaging and action should account for *why* these populations are considered hidden. While used for surveillance, this sampling strategy also has implications for ongoing mpox research and response in DRC and similar low-resource contexts where the disease is endemic.

## Data Availability

Protocols and questionnaires used in this study will be shared upon reasonable request to the authors.

## ACKNOWLEDGEMENTS

We would like to acknowledge the support of our IMReC collaborators including: Muge Cevik, Souradet Shaw, Issac Bogoch and Gregg Gonsalves in the creation of this study. We would also like to acknowledge the peer educators in Kinshasa, Kenge, and Goma who worked to recruit these hidden populations. Without their involvement, this work would not have been possible. Figures for this manuscript were created using Biorender.com.

## FUNDING

This work was supported by the United States Department for Agriculture (USDA) Agriculture Research Service (ARS) (USDA ARS NACA number 20230048, grant number 58-3022-2-020) and by the International Mpox Research Consortium (IMReC) through funding from the Canadian Institutes of Health Research and International Development Research Centre (grant no. MRR-184813). These findings do not represent official views or endorsements of the United States or Canadian governments.

## DATA SHARING

Protocols and questionnaires used in this study can be shared upon reasonable request to the authors.

## Notes

### Competing Interest Statement

The authors have declared no competing interest.

### Author Declarations

This study was approved by the Institutional Review Boards of the University of California, Los Angeles (IRB#23-000676), University of Manitoba (HS25837) and the Kinshasa School of Public Health, University of Kinshasa, DRC (ESP/CE/161/2024). All participants were provided with an informed consent form which was signed or marked in agreement before commencing any study activities.

